# Topic: Effect of Lockdown Of COVID-19 And The Impact On People Living In Enugu State, Nigeria

**DOI:** 10.1101/2023.11.10.23298376

**Authors:** Nwadiuto C Ojielo, Daniel C Onwuliri, Augustine Onuh, Ada Ilo, Ngozi R Njeze

## Abstract

**Introduction:** When the coronavirus (COVID-19) pandemic struck, countries employed diverse strategies in the control of the virus aimed at preventing, detecting, controlling and mitigating the impact of the pandemic. Initially, there was no known cure for this viral infection so this ‘stay-at-home’ necessitated the preventive measures to break the chain of transmission. This study aims to examine the different challenges people passed through as a result of this pandemic and the effect of the lockdown on people. This study will serve as the basis for future reference to know if this lockdown could be repeated in the face of another pandemic.

**Methodology:** The study was carried out in Enugu Metropolis, South East, Nigeria. This is a cross-sectional study among people living in Enugu State. Google forms were dispatched to adults from 18-65 years on social media or the internet. Data entry was to Excel, then to SPSS version 26 and analyzed with student’s t-test and Chi-square.

**Results:** From the study, 35% said there were financial losses, some complained of poor recreation, 52% of people said there was loss of learning opportunities for their children due to school shutdown 23% had a reduction in their income with 35% losing their job, 57.9% of people said the crime rate increased, 26.6% had a mental breakdown, 45.1% of people had relationship/spousal problems and 64.1% of people lost someone during the COVID-19 lockdown and There were also some benefits of the lockdown like 70% said there was reduction in RTA, air pollution, introduction to remote working popularly known as “working from home” and 32% of people claimed it caused family bonding. In all, 73% of people said they would not want or support a repeat lockdown.

**Conclusion:** The impact of the lockdown on the residents of Enugu State was generally not a palatable experience. From this study, though some good things were achieved, it led to a number of irreversible crises. It also caused serious implications like a decline to access to healthcare, economic effects, political, social and cultural effects, educational impacts, an increase in crime rate, domestic violence, religious impact, and environmental effects. In future, a better method of approach is recommended.

## Introduction

Coronavirus disease 2019, abbreviated to COVID-19, is a viral infectious disease caused by severe acute respiratory syndrome coronavirus 2 (SARS-CoV-2) which is a member of the Corona viridae family. One unique characteristic of this virus is its ability to spread rapidly from a single city to the entire country within a short period.^1^ The speed of both the geographical expansion and the sudden increase in the number of cases is capable of overwhelming health services in most countries. The World Health Organization declared it a pandemic and a public health emergency of international concern.^2^

Since the coronavirus disease (COVID-19) pandemic, countries have continued to employ diverse strategies in the control of the virus. These strategies are aimed at preventing, detecting, controlling and mitigating the impact of the pandemic. The initiatives vary in their differential effectiveness, but one approach that has been adopted by several countries is the lockdown strategy. This is a "stay at home measure" to slow the spread of the virus which started in Nigeria on the 30th March 2020.^3^

The President of the Federal Republic of Nigeria, President Muhammadu Buhari issued a lockdown of non-essential activities in Lagos State, the FCT and Ogun State for an initial period of 14 days. The lockdown as a strategy was designed to present a window of opportunity that would enable the Ministry of Health and National Centre For Disease Control(NCDC) to achieve the following in every state in the Country: Enabling prompt detection and treatment of cases, Limiting transmission in areas where cases had been reported, Limiting the spread of new cases to other geographical areas, enhancing contact tracing activities and efficient use of resources and Improving sample collection (NCDC 2021).^4^

Initially, there was no known cure for this viral infection at that time. Therefore, this necessitated preventive measures to break the chain of transmission. To limit the spread of the disease, people were forced to stay at home. However, this promising preventive measure hurt the health of the people as well as other aspects of their lives such as work, academics religion and even society. The psychological effect of the stay-at-home order has been underestimated. Also, more attention was shifted to COVID-19, while people with other debilitating ailments like cardiac diseases, endocrine disorders and even cancers were given less attention. Other essential routine health care such as routine immunization were neglected.^5^ People who suffer from respiratory diseases such as asthma or cardiac failure were mistaken for COVID-19 and were not given adequate attention. Resources for other health system strengthening were diverted to COVID-19 whereas health workers became apprehensive of managing and caring for sick people due to fear of contracting the deadly virus.

This condition impacted on sexual and reproductive health; with a drastic reduction of reproductive health services globally, an infringement on the rights of women and girls. The United Nations has raised fear about the expected impact of COVID-19 on women and girls in the background of their special health needs such as maternal health services, menstrual hygiene and family planning services.^6^ Therefore, women and girls are said to be more vulnerable in times of health crises like COVID-19 and are usually denied access to essential commodities such as family planning, menstrual pads and Antenatal care at times like this exposing them to exacerbation of maternal morbidity and mortality. In Latin America and the Caribbean for instance, 18 million women have lost access to regular modern contraceptives.^7^ Also, most women lost their jobs because they had to take care of their children who stopped going to school due to the sit-at-home.

On the other hand, women are at risk due to their number in the workforce. It has been stated that women make up seventy percent of the workforce and are also more likely to be the frontline health workers in many countries especially nurses, community health workers and midwives.^8^ There was a paucity of academic studies that could guide researchers to study the impact of this pandemic on the health and economies of countries.

Therefore, this study aims to examine the different challenges people passed through as a result of this pandemic and the effect of the lockdown on people. This study will serve as the basis for future reference to know if this lockdown could be repeated in the face of another pandemic.

## Methodology

### Study Area

The study was carried out in Enugu Metropolis, South East, Nigeria. The metropolis is made up of three LGAs in Enugu State (Enugu North, Enugu South and Enugu East) and is inhabited mostly by Ibo-speaking people. Enugu state had a population of 1,596,042 males and 1,671,795 females.^9^ The state shares borders with Abia State and Imo State to the south, Ebonyi State to the east, Benue State to the northeast, Kogi State to the northwest and Anambra State to the west.

The state has three senatorial zones Enugu East, Enugu West and Enugu North but the metropolis is in Enugu East zone. The state is noted for its coal deposit, the largest in Africa, giving it its name Coal City State.

### Study Design

This is a cross-sectional study of people living in Enugu State who owned smartphones. Google forms were dispatched to adults from 18-65 years on social media or the internet. Government workers and very ill patients were excluded. The filled forms were sent back through social media and subsequently analyzed.

A total of 467 Google forms were sent, after considering inclusion and exclusion criteria. 304 people responded (filled out the forms and sent them back). The duration of this study was 3 months. Data collected started on the 8^th^ of December, 2020 and ended on the 14^th^ of March, 2021.

### Data Collection

A total of 467 Google forms were sent to people with smartphones. Three hundred and four (304) people filled out the forms and sent them back.

### Data Analysis

Data was entered to Excel and then to SPSS version 26. Quantitative variables were summarized using means and standard deviations and analysed with a student’s t-test and paired t-test. Categorical variables were summarised using frequencies and percentages.

### Ethics declarations

The study and experimental protocols were approved by the Ethics Committee of the University of Nigeria Teaching Hospital, Enugu State, Nigeria on 7th December 2020 (NHREC/05/01/2008B-FWA00002458-1RB00002323). Informed consent was obtained from all individual participants included in the study. All procedures performed in studies involving human participants were by the ethical standards of the Ethics Commission of the University of Nigeria Teaching Hospital and with the 1964 Helsinki Declaration and its later amendments or comparable ethical standards.

Consent was written which was documented and sent to the participants before filling the questionnaire forms. They were explained the need to withdraw at any time with no consequence. This study did not include minors.

## Results

**Table 1.** -Sociodemographic characteristics. From the sociodemographics, majority of the respondents (60%) were females and between the ages of 31 to 40 years (43%). A high number of them (82.6%) were married and most of them were medical doctors (45.7%), Christians (92.4%) and from Ibo (80.26%).

| **Socio-demography** | ***Frequency***<br>**(N=304)** | ***Percentage***<br>**(%)** |
| --- | --- | --- |
| **Gender** |  |  |
| Male | 121 | 40 |
| Female | 183 | 60 |
| **Age** |  |  |
| 11 to 20 years | 33 | 11 |
| 21 to 30 years | 64 | 21 |
| 31 to 40 years | 131 | 43 |
| 41 to 50 years | 49 | 16 |
| 51 to 60 years | 21 | 7 |
| Above 60 years | 6 | 2 |
| **Marital status** |  |  |
| Single | 53 | 17.4 |
| Married | 251 | 82.6 |
| **Occupation** |  |  |
| Medical doctor | 139 | 45.7 |
| Business / traders | 85 | 28 |
| Students | 47 | 15.46 |
| Others | 33 | 10.86 |
| **Religion** |  |  |
| Christianity | 281 | 92.4 |
| African traditional religion | 1 | 0.33 |
| Islam | 22 | 7.24 |
| **Ethnicity** |  |  |
| Igbo | 244 | 80.26 |
| Yoruba | 18 | 5.92 |
| Hausa | 11 | 3.62 |
| Others | 31 | 10.2 |

**Table 2.**
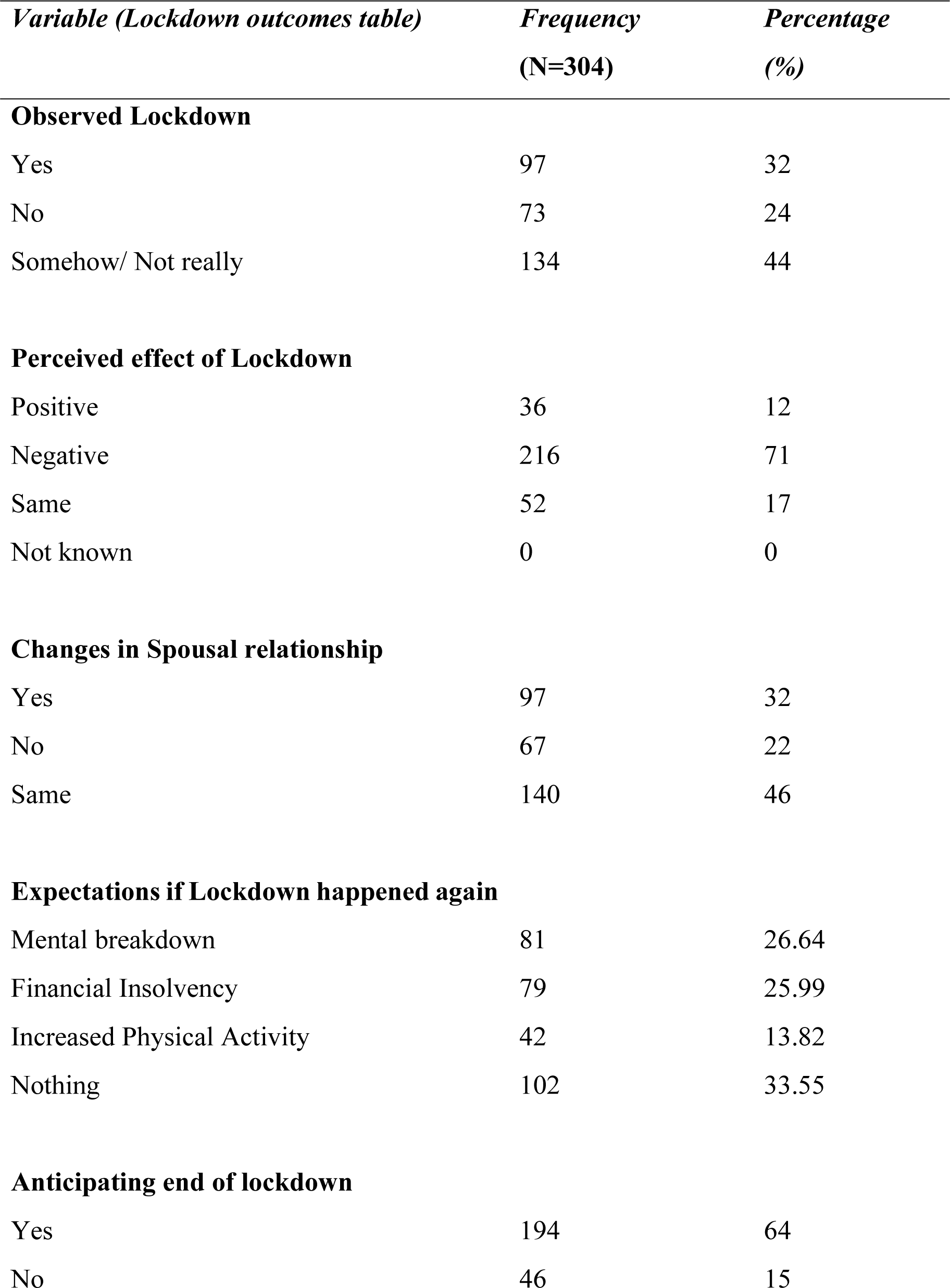
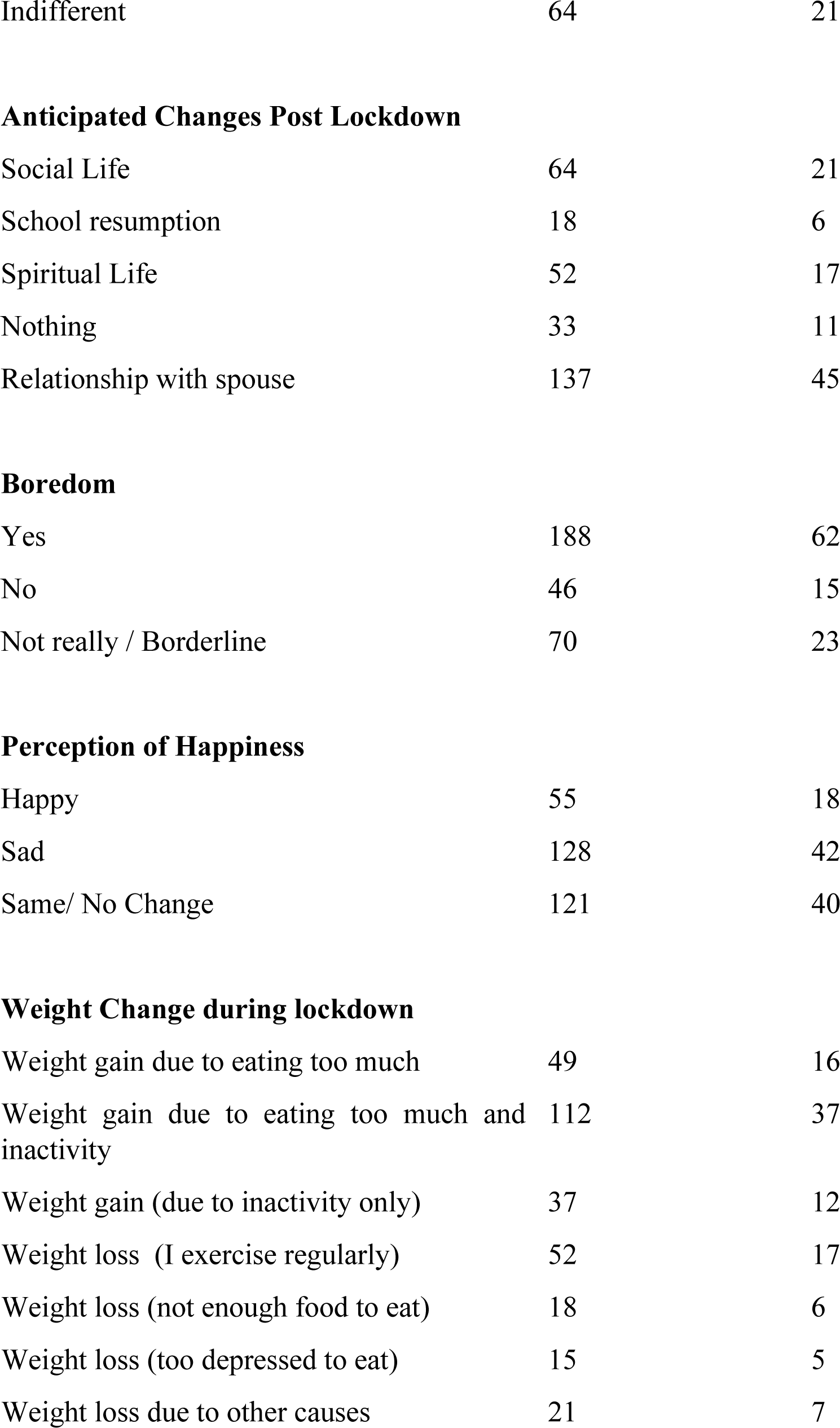
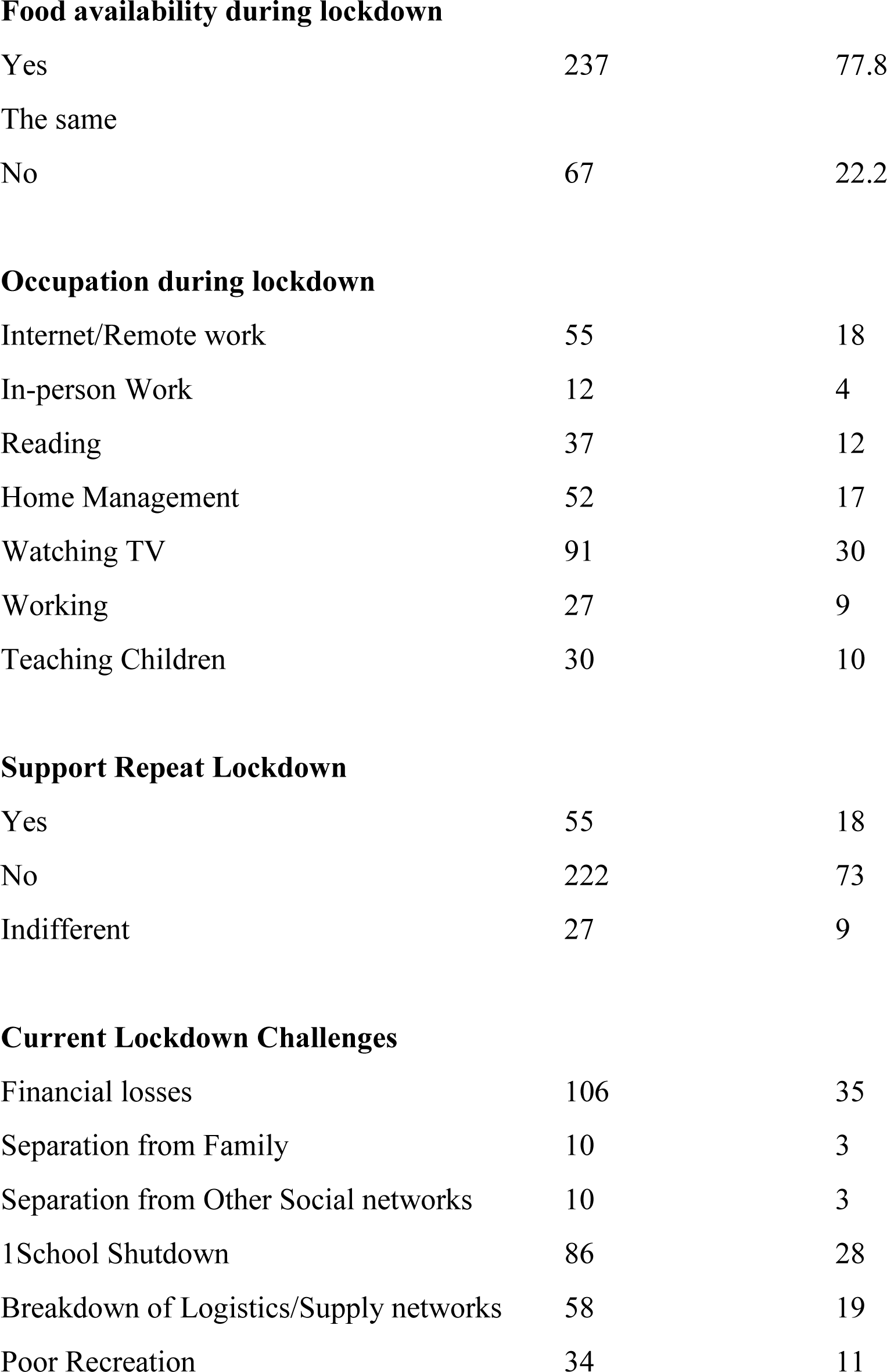
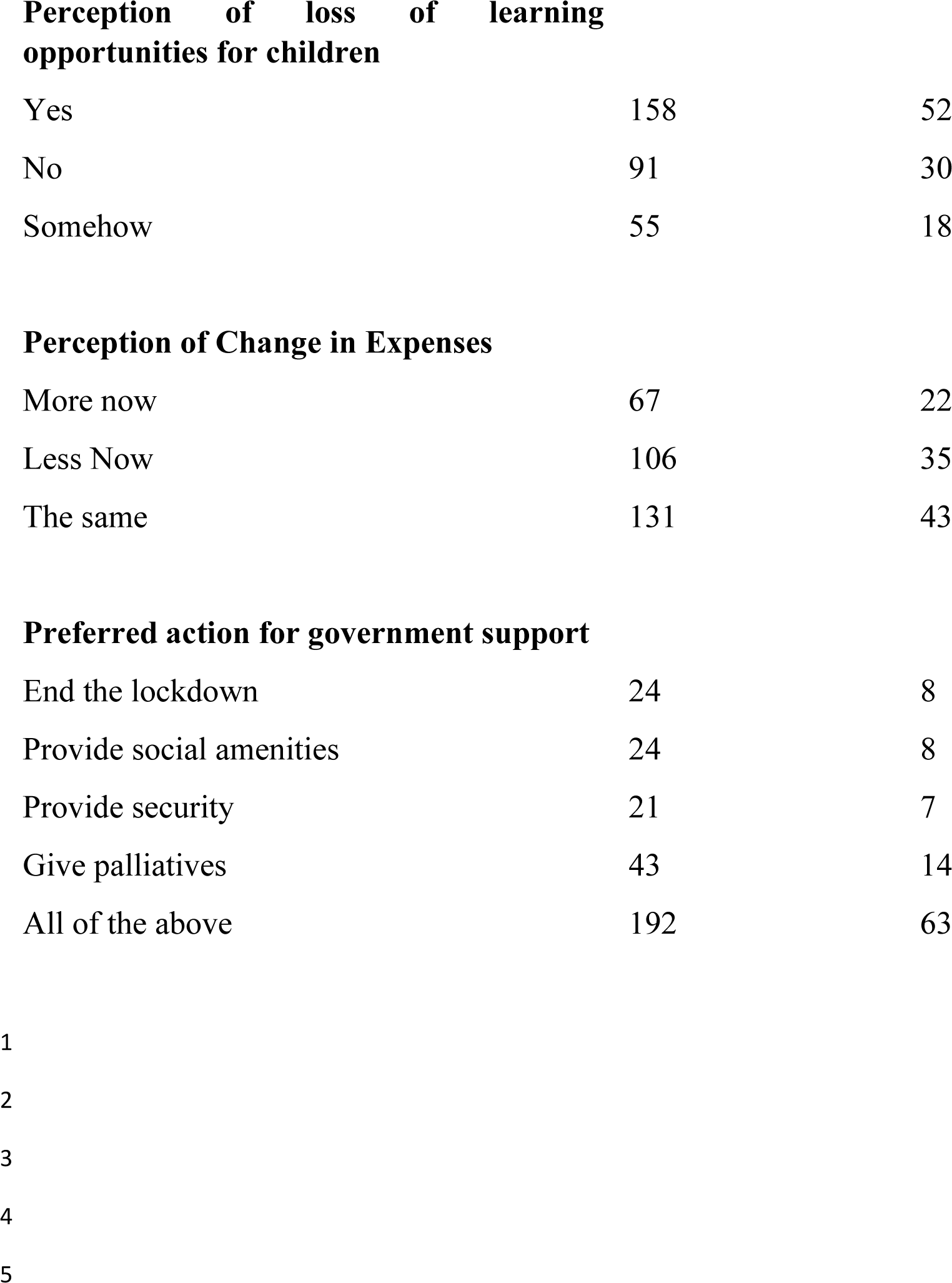
– Effects of lockdown. Table 2 shows the perception of the lockdown, majority (62%) were bored, watching television (30%) and a lot gained weight (37%). Most perception about the lockdown was negative (71%) and the different challenges faced during the lockdown

| *Variable (Lockdown outcomes table)* | *Frequency*<br>*(N=304)* | *Percentage*<br>*(%)* |
| --- | --- | --- |
| **Observed Lockdown** |  |  |
| Yes | 97 | 32 |
| No | 73 | 24 |
| Somehow/ Not really | 134 | 44 |
| **Perceived effect of Lockdown** |  |  |
| Positive | 36 | 12 |
| Negative | 216 | 71 |
| Same | 52 | 17 |
| Not known | 0 | 0 |
| **Changes in Spousal relationship** |  |  |
| Yes | 97 | 32 |
| No | 67 | 22 |
| Same | 140 | 46 |
| **Expectations if Lockdown happened again** |  |  |
| Mental breakdown | 81 | 26.64 |
| Financial Insolvency | 79 | 25.99 |
| Increased Physical Activity | 42 | 13.82 |
| Nothing | 102 | 33.55 |
| **Anticipating end of lockdown** |  |  |
| Yes | 194 | 64 |
| No | 46 | 15 |
| Indifferent | 64 | 21 |
| **Anticipated Changes Post Lockdown** |  |  |
| Social Life | 64 | 21 |
| School resumption | 18 | 6 |
| Spiritual Life | 52 | 17 |
| Nothing | 33 | 11 |
| Relationship with spouse | 137 | 45 |
| **Boredom** |  |  |
| Yes | 188 | 62 |
| No | 46 | 15 |
| Not really / Borderline | 70 | 23 |
| **Perception of Happiness** |  |  |
| Happy | 55 | 18 |
| Sad | 128 | 42 |
| Same/ No Change | 121 | 40 |
| **Weight Change during lockdown** |  |  |
| Weight gain due to eating too much | 49 | 16 |
| Weight gain due to eating too much and inactivity | 112 | 37 |
| Weight gain (due to inactivity only) | 37 | 12 |
| Weight loss (I exercise regularly) | 52 | 17 |
| Weight loss (not enough food to eat) | 18 | 6 |
| Weight loss (too depressed to eat) | 15 | 5 |
| Weight loss due to other causes | 21 | 7 |
| **Food availability during lockdown** |  |  |
| Yes | 237 | 77.8 |
| The same |  |  |
| No | 67 | 22.2 |
| **Occupation during lockdown** |  |  |
| Internet/Remote work | 55 | 18 |
| In-person Work | 12 | 4 |
| Reading | 37 | 12 |
| Home Management | 52 | 17 |
| Watching TV | 91 | 30 |
| Working | 27 | 9 |
| Teaching Children | 30 | 10 |
| **Support Repeat Lockdown** |  |  |
| Yes | 55 | 18 |
| No | 222 | 73 |
| Indifferent | 27 | 9 |
| **Current Lockdown Challenges** |  |  |
| Financial losses | 106 | 35 |
| Separation from Family | 10 | 3 |
| Separation from Other Social networks | 10 | 3 |
| 1 School Shutdown | 86 | 28 |
| Breakdown of Logistics/Supply networks | 58 | 19 |
| Poor Recreation | 34 | 11 |
| **Perception of loss of learning opportunities for children** |  |  |
| Yes | 158 | 52 |
| No | 91 | 30 |
| Somehow | 55 | 18 |
| **Perception of Change in Expenses** |  |  |
| More now | 67 | 22 |
| Less Now | 106 | 35 |
| The same | 131 | 43 |
| **Preferred action for government support** |  |  |
| End the lockdown | 24 | 8 |
| Provide social amenities | 24 | 8 |
| Provide security | 21 | 7 |
| Give palliatives | 43 | 14 |
| All of the above | 192 | 63 |

**Table 3.**
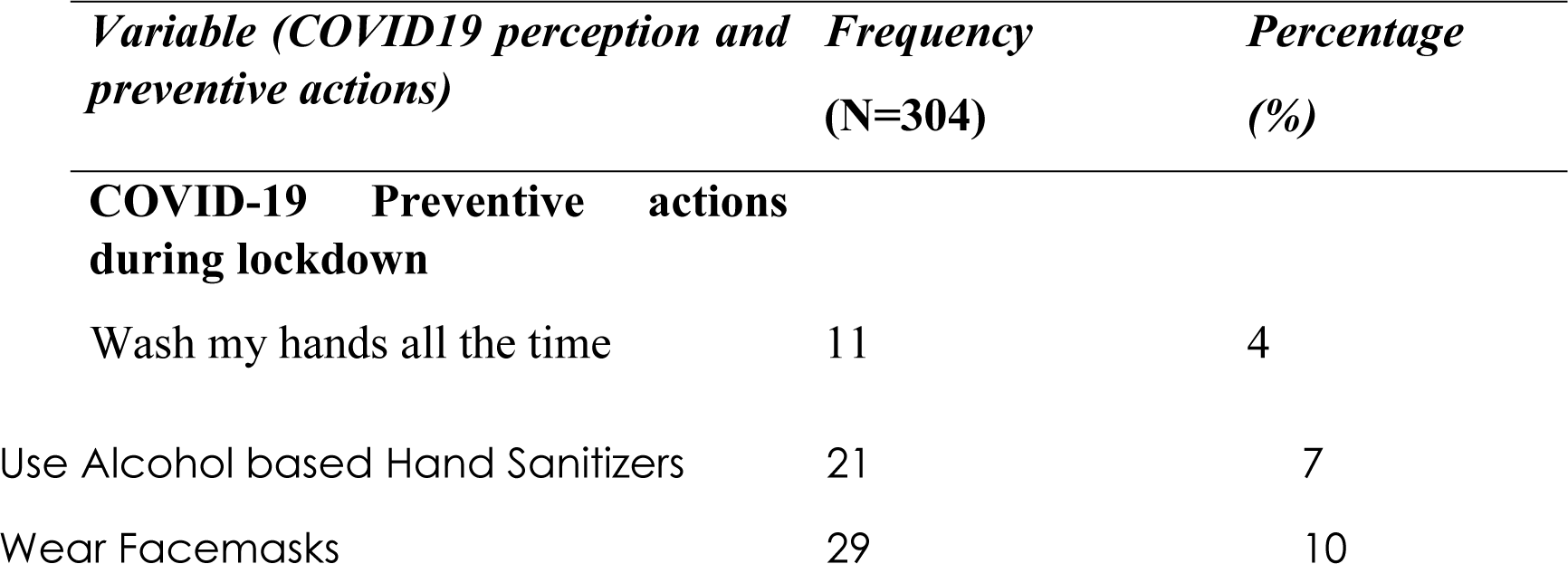
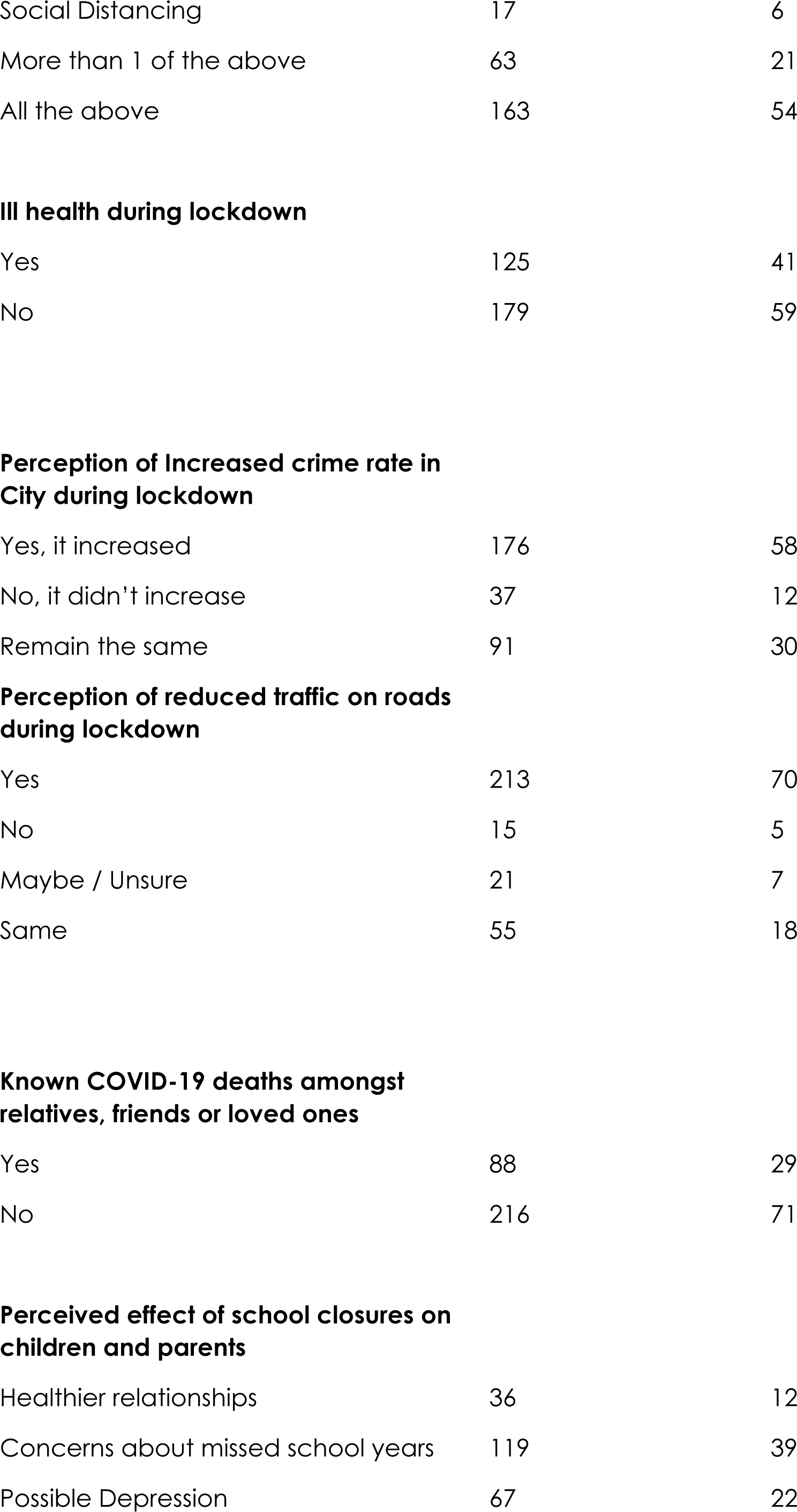
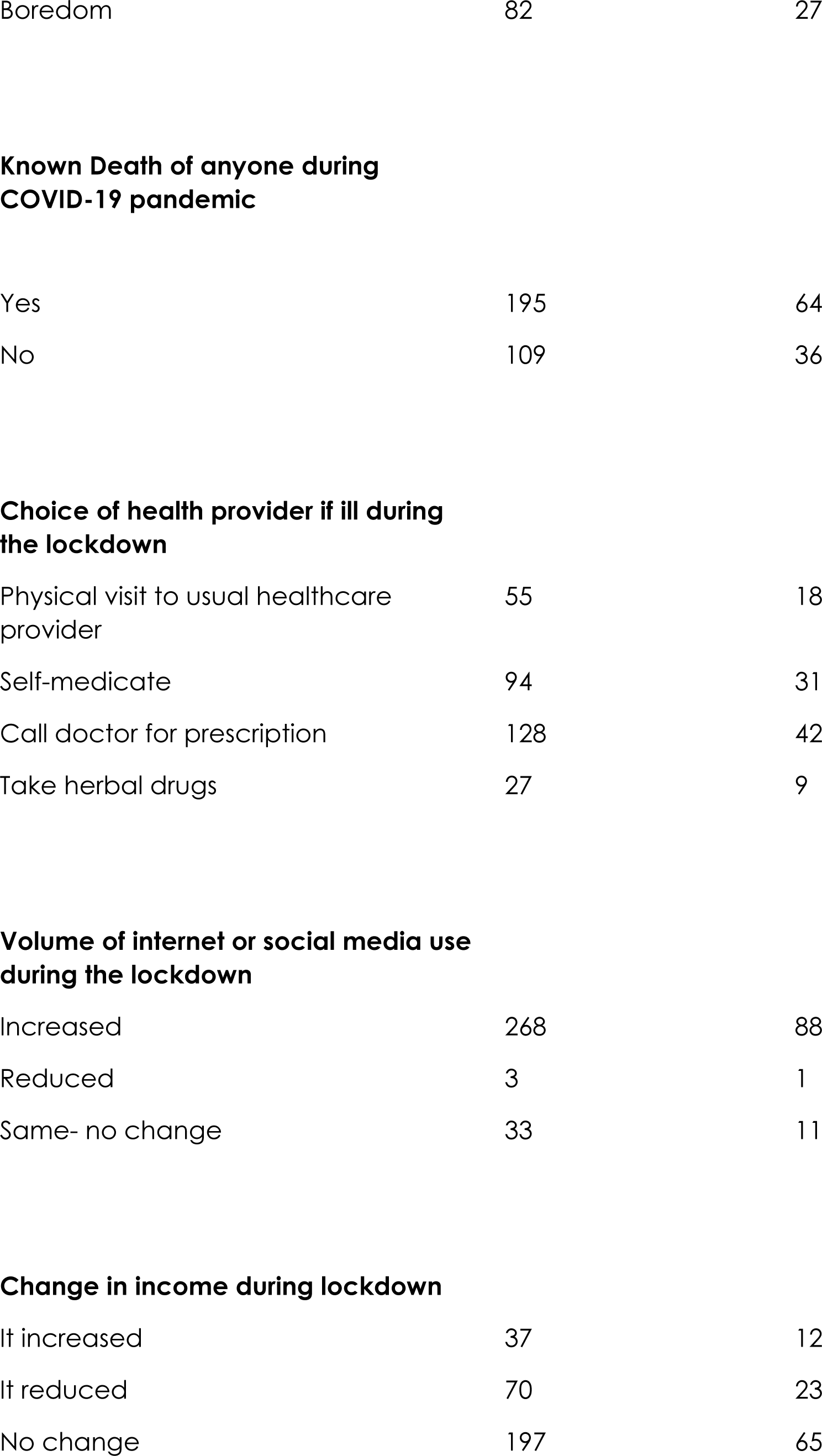
-Effects of the lockdown. 54% of the respondents used both face masks, hand sanitizers and social distancing, in addition to the lockdown to prevent COVID-19. Internet and social media increased (88%) and 58% of people believed there was an increase in crime rate and 23% said they had reduced income. Majority of people (64%) knew someone that died during this lockdown and 41% got sick while 42% called their doctors for prescription when they were sick. However, a lot of people(70%) acknowledged that there was a reduction in RTAs.

| *Variable (COVID19 perception and preventive actions)* | *Frequency*<br>*(N=304)* | *Percentage*<br>*(%)* |
| --- | --- | --- |
| **COVID-19 Preventive actions during lockdown** |  |  |
| Wash my hands all the time | 11 | 4 |
| Use Alcohol based Hand Sanitizers | 21 | 7 |
| Wear Facemasks | 29 | 10 |
| Social Distancing | 17 | 6 |
| More than 1 of the above | 63 | 21 |
| All the above | 163 | 54 |
| **Ill health during lockdown** |  |  |
| Yes | 125 | 41 |
| No | 179 | 59 |
| **Perception of Increased crime rate in City during lockdown** |  |  |
| Yes, it increased | 176 | 58 |
| No, it didn't increase | 37 | 12 |
| Remain the same | 91 | 30 |
| **Perception of reduced traffic on roads during lockdown** |  |  |
| Yes | 213 | 70 |
| No | 15 | 5 |
| Maybe / Unsure | 21 | 7 |
| Same | 55 | 18 |
| **Known COVID-19 deaths amongst relatives, friends or loved ones** |  |  |
| Yes | 88 | 29 |
| No | 216 | 71 |
| **Perceived effect of school closures on children and parents** |  |  |
| Healthier relationships | 36 | 12 |
| Concerns about missed school years | 119 | 39 |
| Possible Depression | 67 | 22 |
| Boredom | 82 | 27 |
| **Known Death of anyone during COVID-19 pandemic** |  |  |
| Yes | 195 | 64 |
| No | 109 | 36 |
| **Choice of health provider if ill during the lockdown** |  |  |
| Physical visit to usual healthcare provider | 55 | 18 |
| Self-medicate | 94 | 31 |
| Call doctor for prescription | 128 | 42 |
| Take herbal drugs | 27 | 9 |
| **Volume of internet or social media use during the lockdown** |  |  |
| Increased | 268 | 88 |
| Reduced | 3 | 1 |
| Same- no change | 33 | 11 |
| It increased | 37 | 12 |
| It reduced | 70 | 23 |
| No change | 197 | 65 |

**Figure 1:**
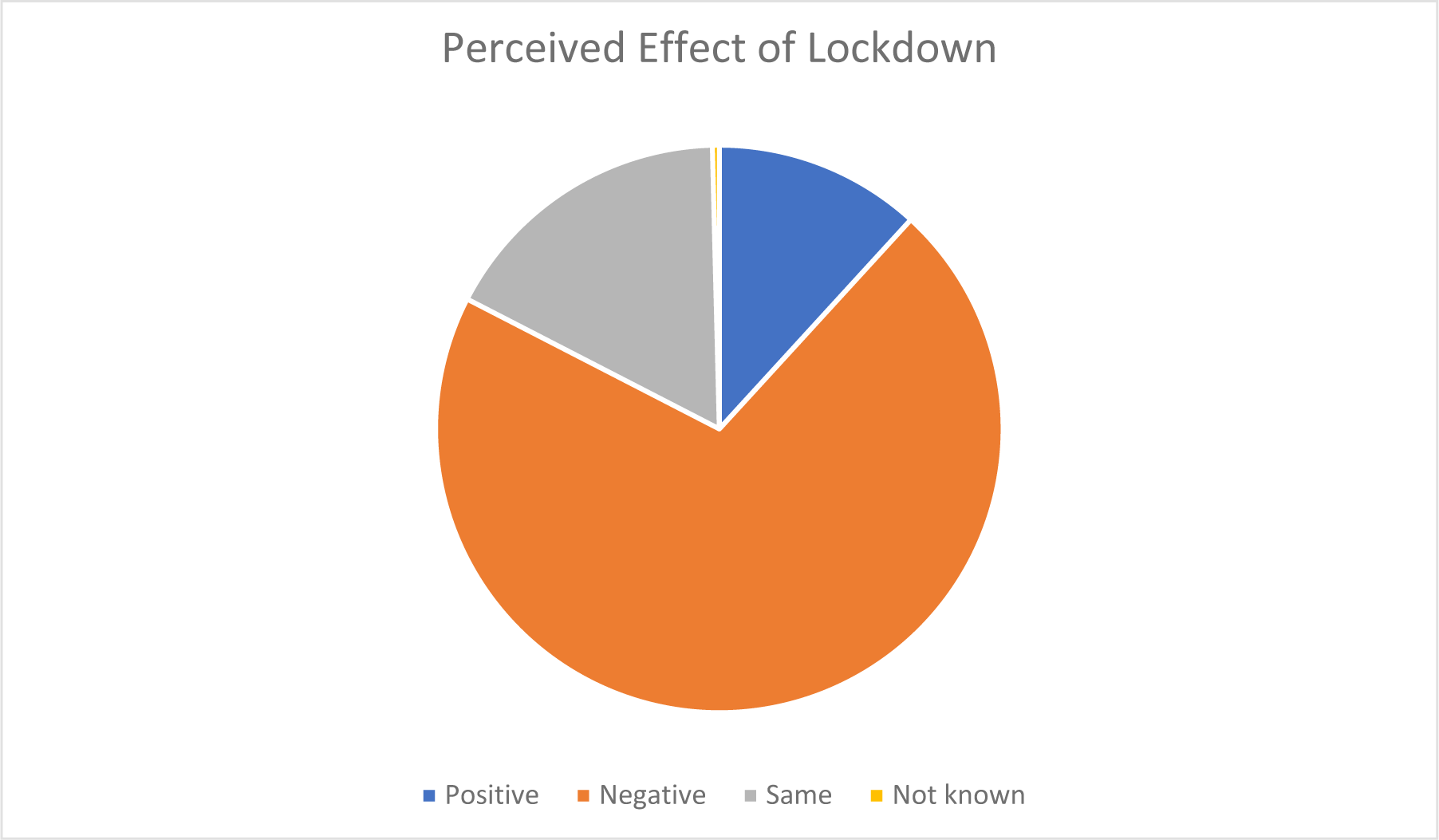
Perceived effect of lockdown. Majority (71%) of people believed the effect of lockdown had a negative impact on them, some believed that there was no effect (17%) while others(12%) believed it had a positive effect on them. A very small amount was undecided

## Discussion

The lockdown of cities and countries due to the coronavirus pandemic impacted every aspect of society, and the healthcare system was not exempted.

The lockdown and quarantine that followed this global disease took a toll on the mental health of the populace. The psychologic and psychiatric disturbances include anxiety, post-traumatic stress disorder, stigmatization, depression, panic attacks, and behavioural changes.

From our analysis, only 32% of respondents observed the lockdown fully 24% said they did not while 44%% said they observed the lockdown partially.

The effects the coronavirus pandemic has had on Africa’s largest economy are starting to show in data. The Nigerian Bureau of Statistics showed that Nigeria’s economy contracted by 6.1% year on year in the second quarter of this year. The dip follows thirteen quarters of positive but low growth rates. The −6.1% decline is also Nigeria’s steepest in the last 10 years.^7^

The COVID-19 pandemic has had far-reaching consequences beyond the spread of the disease itself and efforts to quarantine it, including political, cultural, and social implications, including world peace, education, religion and social issues such as domestic violence and inequality.

The virus has also had an unprecedented impact on the environment. Some studies estimated a positive indirect impact on the environment. Climate experts anticipated that greenhouse gas (GHG) emissions could drop so low to proportions never before seen since World War II.^10^ This result is mostly due to the social distancing policies directed by the governments due to the onset of the pandemic.

An analysis of 22 hospitals in France showed a decrease in all emergency department visits by 26 percent, including a decrease in strokes, transitory ischemic attacks, unstable angina, appendicitis, and seizures.^11^ These events are unlikely to have reduced within the population, which leaves the authors with the conclusion that these events are occurring with no emergency medical intervention.

Other health challenges besides COVID-19 require treatment even in the wake of the pandemic. One such disease that requires ongoing treatment, follow-up and procedures is cancer. In a study published in Target Oncology, it was concluded that contracting the COVID-19 infection along with the breakdown of healthcare, and the economic crisis, will have an unprecedented impact on cancer patients worldwide.^12^

From our study, 41% of people got sick during this lockdown, 29% had relatives and loved ones who died due to Covid-19 and 64% of the people they know died during this period due to both Covid-19 and other causes. 31% of people self-medicated, and 42% of people called their doctors. This showed there were reduced hospital visits (walk-in) from non-communicable diseases during the lockdown.

A recent report from the group that has worked on COVID-19 models at Imperial College London has shown that if all malaria-control activities are highly disrupted, the malaria burden for 2020 could more than double that of 2019, resulting in large malaria epidemics across Africa.^13^ The prevalence of non-communicable diseases during the pandemic was 39.9% as against 29%.^13^

The closure or slowdown in normal clinical services further aggravated the underlying conditions of patients, leading to more severe cases of non-communicable diseases. The highly contagious measles cases continued to increase in Nigeria (Cases in December 2020 were 9,316 compared to 2,064 confirmed cases in November 2019.^5^

A paper published in the Lancet has modelled three scenarios in which coverage of basic life-saving interventions for maternal and child health is reduced for different durations as a result of a combination of factors including reduction in demand for health services, availability of supplies and equipment, as well as health workers.

These early estimates show a worrying 9.8 percent to 44.7 percent increase in under-five child deaths per month and an 8.3 percent to 38.6 percent increase in maternal deaths per month.^14^ Presently, due to the restrictions on movement and lockdown, commuting to the healthcare facilities when children are sick might be delayed.

One of the routine health services that is being disrupted by COVID-19 is childhood immunization. According to the World Health Organization, an estimated 80 million children in 68 countries are at risk of developing vaccine-preventable diseases such as measles, diphtheria, and polio, because of the disruption of routine immunization services.^15^

There was also a decrease in the utilization of services. Immunization was adversely affected by the coming of COVID-19 with a decline in several fully immunized children. Comparing the pentavalent performance in 2019 with that of 2020, it shows a decline in performance especially in the COVID-19 months. ^15^

Another study reported widespread fear, of getting turned away from hospitals, delays in infant immunization, missed immunization opportunities and increasing recourse to traditional treatments as the pandemic spread in Nigeria. There has also been a perceived low quality of care among patients during this period.^7^

Increase in endemic diseases because of the focus on the pandemic, many clinical activities have either been halted or reduced to curb COVID-19 transmission. The number of antenatal care services performed also dropped and by January 2021, it was 2.9% compared to 56.8% as of 2018 (NDHS); the number of Postnatal care services performed-was 7.7% by January 2021, compared to 67.0% in 2018.^4^

Wang and colleagues investigated the psychological impact of the pandemic, including anxiety, depression, and stress during the initial stage of the outbreak in China.^16^ COVID-19 has already been recognized as a cause of direct and indirect psychological and social consequences that might impact mental health not only during the pandemic per se but also in the future.^17^ Indeed, the quarantine effects have already been explored during past outbreaks, such as during the outbreak of Severe Acute Respiratory Syndrome (SARS) in 2003 and Ebola in 2014, indicating that the mental health impact can be broad, massive and long-lasting. Quarantine can be very stressful; period, especially if one is away from loved ones. Among the consequences of quarantine, are acute stress disorders, anxiety, irritability, poor concentration and indecisiveness, deteriorating work performance, post-traumatic stress disorders, high psychological distress, depressive symptoms and insomnia.

The lockdown was particularly oppressive and caused financial insecurity and subsequently depression because merchants were hoarding their wares as a result of fear of not finding goods to buy and sell. And many people lost their jobs, and as expected depression would arise from this severe financial uncertainty. From our study, 35% of respondents had a financial loss The effect on drug addicts was also enormous as they were compelled to stay indoors; so they had withdrawal syndrome since they could not access their illicit drug. It is not surprising that the pandemic and concomitant lockdown are presenting a remarkable increase in psychiatric disorders in health workers and the general public.^17^

The researchers suggest that restrictions on movement and fears about COVID-19 transmission might have prevented caregivers from accessing services. In addition to this, the negative impact of the lockdown on the economy has affected the cash reserves of families, rendering them unable to afford healthcare, as the major means of healthcare financing in Nigeria is out-of-pocket payment.

Community participation is essential in the collective response to coronavirus disease 2019 (COVID-19), from compliance with lockdown to the steps that need to be taken as countries ease restrictions, to community support through volunteering. Unfortunately, this has largely not been the case with Nigeria. A large number of people have not cooperated with the guidelines due to a lack of sustenance and livelihood.^12^

Healthcare workers were impacted physically and psychologically; the lockdown does not apply to them as they are essential workers and are indispensable in the battle against coronavirus disease, but quarantine or self-isolation may be necessary. Healthcare workers are more vulnerable to COVID-19 infection than the general population due to frequent contact with infected individuals. Healthcare workers have been required to work under stressful conditions without proper protective equipment and make difficult decisions involving ethical implications.^15^ This breeds frustration. An Italian nurse committed suicide after being traumatized trying to save the lives of those with COVID-19.20 This emphasizes the need for better working conditions for healthcare workers and closer attention to their mental health.

The lockdown has also negatively affected the training of healthcare personnel. Undergraduate medical training particularly in Nigeria has been put on hold as universities closed down. For post-graduate medical training, there have been delays such as postponed examinations and workshops. The West African and National Postgraduate Medical Examinations all failed to be held in April/May 2020. Local and international conferences, trainings, workshops and seminars have been delayed or cancelled altogether, thereby putting a strain on medical training. Hospitals suffered from the decline in other patients. The private healthcare sector witnessed an 80% fall in patient visits, leading to salaries of clinical staff being reduced or frozen, and some staff being furloughed or even laid off.^18^

Black people are among the worst affected by the COVID-19 epidemic unlike in the Western countries where the spending is less out-of-pocket. Healthcare in Nigeria is largely financed by out-of-pocket spending.

A persistent and major weakness of the country’s health system is the poor functioning of the health financing building block, which is characterized by low public spending, very high levels of out-pocket spending (one of the highest in the world), high incidence of catastrophic health spending and impoverishment due to spending on healthcare. With the worsening of the economy during the lockdown, the state of health financing in Nigeria is catastrophic indeed. Increased hospital bills due to additional PPE charges and low accessibility of cash during the period of the lockdown further worsened the condition.

The lockdown in Nigeria and its management was a policy decision of the federal government in conjunction with the state governments. The enforcement was however poorly coordinated and sometimes subject to the whims and caprices of the security agencies. Some healthcare workers were at various times and in various places harassed by security agencies on their way to or from work, while some patients were turned away or unduly delayed on their way to access healthcare. From our study, 63% of respondents expected government intervention by giving palliatives, providing security, providing social amenities and if possible, ending the lockdown.

The supply of essential medicines and supplies was threatened by the lockdown. This includes antibiotics, vaccines, consumables like gloves, masks, and resuscitation fluids. This is felt most acutely in developing countries. The COVID-19 pandemic triggered several challenges that have led to shortages and price hikes, and could potentially fuel an epidemic of fake and substandard medicines.^19^

The surge in demand and pressure on supply chains has also resulted in severe constraints on the supply of raw materials, especially for N95 masks, surgical masks, and medical gowns, which are the products in highest demand (given their importance for frontline healthcare workers). This has led to a shortage and therefore increase in the costs of PPE.

Ventilators are essential to support lung function, especially for patients with the most severe cases of COVID-19 infection. As the COVID-19 pandemic spread, the demand for ventilators increased worldwide. With limited stock, governments and health systems around the world enlisted the help of various companies to produce medical ventilators. 100 more ventilators were purchased (in addition to the existing 350), although these too, were not enough, considering the growing caseloads.^4^

Maintenance of equipment already owned by healthcare centres was also put on hold or difficult to access. Medical Equipment maintenance in Nigeria took a hit during the pandemic due to restricted local and international travel. The spares and technical know-how for the maintenance of most medical devices, normally scarce in Nigeria were exacerbated by the restrictions of the lockdown.

It is estimated that during the lockdown periods Nigeria’s GDP suffered a 34.1 percent loss due to COVID-19, amounting to USD 16 billion, with two-thirds of the losses coming from the services sector.^20^ The agriculture sector, which serves as the primary means of livelihood for most Nigerians, suffered a 13.1 percent loss in output (USD 1.2 billion). As with most other economies around the world, the sharp drop in Nigeria’s GDP growth is largely down to the slowdown in economic activity after the country resorted to a lockdown back in April to curb the spread of the virus. In the wake of the pandemic, the World Bank forecast a decline of −3.2% for 2020—a five percentage point drop from its previous projections.^20^

Furlough and layoff of staff employment. Salaries were slashed. Businesses were closed. The aviation sector suffered losses. Banks closed branches. The service sector was the hardest hit.

However, some sectors saw a boom in sales (Pharmaceuticals, Medical consumables specifically Hand sanitisers, soap and detergents, plastic industries, chemical markets, home delivery services like Amazon). Rice boom (huge sales due to bulk buying).

Emergency Palliatives Committee (EPC) was set up as a COVID relief strategy for the masses, a bailout for businesses and food support for the people.

Nigeria’s economy has also been crippled by external factors such as the near-total shutdown of economic activity around the world during the lockdown. The accompanying steep drop in oil prices amid a drop in global demand left Nigeria drastically short of earnings given its dependence on the commodity as its biggest revenue source. For context, the United States slashed its Nigerian crude oil imports oil by 11.67 million barrels in the first five months of 2020, compared to what it bought in the same period of 2019. In fact, in the second quarter of 2020, local oil production dropped to its lowest since 2016—when Nigeria endured a full year of negative growth.^21^

The latest economic data shows Nigeria’s government continues to fall far short of projections in its Economic Recovery and Growth Plan, created in the aftermath of the 2016 recession. World Bank predicts Africa’s most populous country is set for its worst recession in four decades.

From this study, 22.2% of respondents said there was not enough food available during the lockdown. However, 37% of people gained weight due to inactivity and eating too much, out of boredom, even though 6% of people claimed they lost weight due to lack of food during the lockdown.

22% of respondents said they were spending more during the lockdown .88% of participants believed internet usage increased during this lockdown. 54% of people observed all the infection prevention protocols like the use of hand sanitisers, and face masks, washing hands frequently and maintaining social distancing.

We estimate that households lost on average 33 percent of their incomes during the period, with the heaviest losses occurring for rural non-farming and urban households. This study shows that 23% of respondents said their income/source of income was reduced during this lockdown period and 35% of respondents had a financial loss.

The coronavirus pandemic appears to have worsened conflict dynamics. It has also led to a United Nations Security Council resolution demanding a global ceasefire. On March 23, 2020, United Nations Secretary-General Antonio Guterres issued an appeal for a global ceasefire as part of the United Nations’ response to the COVID-19 coronavirus pandemic and calling for greater international cooperation to address the pandemic.^22^

According to data released by UNESCO on 25 March, school and university closures due to COVID-19 were implemented nationwide in 165 countries. Including localized closures, this affects over 1.5 billion students worldwide, accounting for 87% of enrolled learners.30. From this study, 30% of respondents occupied their time with watching television. 52% of respondents thought their kids were missing out on school, even with internet schooling and 39% of respondents felt that their children missing school would hurt them like depression and increase in crime rate.

Churches were forced to close because of the coronavirus. The pandemic impacted religion in various ways, including the cancellation of the worship services of various faiths, as well as the cancellation of pilgrimages surrounding observances and festivals. Many churches, synagogues, mosques, and temples have offered worship through livestream amidst the pandemic. Relief wings of religious organisations have dispatched medical supplies and other aid to affected areas. In the United States, Trump designated 15 March 2020 as a National Day of Prayer for "God’s healing hand to be placed on the people of our Nation".^23^

The impact on personal gatherings was strong as medical experts advised, and local authorities often mandated stay-at-home orders to prevent gatherings of any size, not just the larger events that were initially restricted. Such gatherings may be replaced by teleconferencing, or in some cases with unconventional attempts to maintain social distancing with activities such as a balcony sing-along for a concert, or a "birthday parade" for a birthday party.

Many countries have reported an increase in domestic violence and intimate partner violence attributed to lockdowns amid the COVID-19 pandemic. Financial insecurity, stress, and uncertainty have led to increased aggression at home, with abusers able to control large amounts of their victims’ daily lives. United Nations Secretary-General António Guterres has called for a domestic violence "ceasefire". From this study, 45% of respondents said there would be a change in their relationships with their spouses by the time the lockdown is over.

Older people are particularly affected by COVID-19. They need special attention during the COVID-19 crisis. While the number of older persons is relatively and smaller in developing countries, particularly in Africa, this coincides with other serious structural risks. Countries with the fewest older persons have the fewest health resources, limited experience caring for older patients (including few geriatric specialists), less institutional care for older persons, and far fewer public or NGO support structures for outreach, screening and community-based care of older persons.

Older persons living in long-term care facilities, such as nursing homes and rehabilitation centres, are particularly vulnerable to infection and adverse outcomes from COVID-19. Older persons who live alone may face barriers to obtaining accurate information, food, medication, and other essential supplies during quarantine conditions and community outreach is required. Older persons, especially those in isolation, those with cognitive decline, and those who are highly care-dependent, need a continuum of practical and emotional support through informal networks (families), health workers, caregivers, and volunteers.

There are Decreased concentrations of NO2 and PM 2.5. 91% of the world’s population live in places where poor air quality exceeds the permissible limits.^24^ The consequences of air quality degradation are demonstrated in a significant percentage of global mortality each year. China implemented strict traffic restrictions and self-quarantine measures to control the spread of SARS-CoV2. These actions generated a change in air pollution. Due to quarantine, NO2 was decreased by 22.8 μg/m3 and 12.9 μg/m3 in Wuhan and China, respectively. PM 2.5 fell by 1.4 μg/m3 in Wuhan but decreased by 18.9 μg/m3 in 367 cities.^25^

Additionally, the Copernicus Atmosphere Monitoring Service (CAMS) of the European Union observed a drop of PM 2.5 last February about the previous three years. In China alone, all of these air quality improvements generated human health benefits that have outnumbered confirmed SARS-CoV2 deaths thus far.^26^

Additionally, 58% of respondents believed that crime increased during this lockdown period but 52% believed that Road traffic accidents reduced due to the lockdown (even though domestic violence increased). However, 70% of respondents observed that traffic, with its effects reduced during this period of lockdown.

Beaches are one of the most important natural capital assets found in coastal areas. They provide services (land, sand, recreation, and tourism) that are critical to the survival of coastal communities and possess intrinsic values that must be protected from over-exploitation.^27^ The lack of tourists, as a result of the social distancing measures, has caused a notable change in the appearance of many beaches in the world. For example, beaches like those of Acapulco (Mexico), Barcelona (Spain), or Salinas (Ecuador) now look cleaner and with crystal clear waters.^28^

Environmental noise is one of the main sources of discomfort for the population and the environment, causing health problems and altering the natural conditions of the ecosystems.

With the enforcement of quarantine measures, the use of private and public transportation has decreased significantly. Also, commercial activities have stopped almost entirely.^29^ These changes have caused noise levels to drop considerably in most cities in the world. 58% of people believed that crime increased during this lockdown period but 52% believed that Road traffic accidents reduced due to the lockdown (even though domestic violence increased). However, 70% of people observed that traffic, with its effects reduced during this period of lockdown.^30,31^

There were also negative impacts on the environment like increased waste. The generation of organic and inorganic waste is indirectly followed by a wide range of environmental issues, such as soil erosion, deforestation, and air, and water pollution.^32^

The quarantine policies, established in most countries, have led consumers to increase their demand for online shopping for home delivery.^33^ Consequently, organic waste generated by households has increased. Also, food purchased online is shipped packed, so inorganic waste has also increased.^34^

Medical waste is also on the rise. Hospitals in Wuhan produced an average of 240 metric tons of medical waste per day during the outbreak, compared to their previous average of fewer than 50 tons. In other countries such as the USA, there has been an increase in garbage from personal protective equipment such as masks and gloves.^33^

Also, there is a reduction in waste recycling. Recycling is a common and effective way to prevent pollution, save energy, and conserve natural resources. As a result of the pandemic, countries such as the USA have stopped recycling programs in some of their cities, as authorities have been concerned about the risk of COVID-19 spreading in recycling centres. In particularly affected European countries, waste management has been restricted. For example, Italy has prohibited infected residents from sorting their waste.^34^ Also, the industry has seized the opportunity to repeal disposable bag bans, even though single-use plastic can still harbour viruses and bacteria.^35^

In Nigeria, the lockdown led to the generation of enormous domestic wastewater and solid waste within the residential areas and along major streets. Wastewater in the form of sewage is seen in gutters, drainage channels, along streets and in open spaces.^36^ Similarly, hospitals, clinics and medical stores during this pandemic have increased amounts of medical waste and infectious waste.^37^ Healthcare workers are encouraged to wear. Personal protective equipment (PPE). Medical waste may be carelessly managed in this critical period of the COVID-19 pandemic.^18^ COVID-19 and Challenges of Management of Infectious Medical Waste in Nigeria). Some of the waste is disposed of at dumpsites or burnt without compliance with the rules stipulated by the National Environmental Standard and Regulation Enforcement Agency (NESREA).^38^

In the same vein, during the lockdown, many less privileged families who cannot afford gas or kerosene as cooking fuel have been cutting down trees in the communities for fuel purposes.^39^ NESREA provided guidelines for handling infectious waste within the context of the pandemic as well as other waste. ^40^

COVID-19 Lockdown has aided the reduction of Air Pollution. A study sought the opinions of 180 residents in the metropolis through the administered questionnaires and found out that 86% agreed that the lockdown has greatly reduced air pollution and greenhouse gas production.^41^

Similarly, an assessment of pollution by the National Aeronautics and Space Administration and the European Space Agency showed that the reduced transport activities have led to limited consumption of energy and less oil demand. These changes have had an important impact on the environmental air quality.^42,43^

It is essential to mention that although the emissions of some Green House Gases (GHG) have decreased as a result of the pandemic, this reduction could have little impact on the total concentration of GHGs that have accumulated in the atmosphere for decades.^44^ For a significant decline, there should be a long-term structural change in the countries’ economies.^45^ This result can be achieved through the ratification of the environmental commitments made. Furthermore, the decrease in GHG emissions currently observed in some countries is only temporary. Once the pandemic ends, countries will most likely revive their economies, and GHG emissions will skyrocket again.^46,47^

Finally, from this study, 71% of people perceived the effect of this lockdown to be negative, with 62% saying they were bored, and 42% being sad. Many of the respondents (35%) had a financial loss and 32% had a negative change in their relationship with their spouse 64% of people were anticipating the end of the lockdown, 27% Of respondents said they would have a mental breakdown if this lockdown happens again. Nevetheless, 73% of people did not support a second/repeat lockdown.

## Conclusion

This Covid-19 pandemic has limited social freedom for all. It also caused serious implications like a decline in access to healthcare, thus causing a rise in non-communicable diseases, mental health, economic effects-increase in financial expenditure and global economic crises, recession, issues of unemployment and "laying-off", political, social and cultural effects, educational impact like closing of schools with increase in crime rate, coronavirus and inequality, increase in domestic violence, religious impact, environmental effects like increased waste, reduction in waste recycling, reductions in greenhouse gas emissions were observed due to significantly reduced road transport, reduced industrial amongst others. In future, a better method of approach is recommended after considering all these.

## Data Availability

All relevant data are within the manuscript and its Supporting Information files.

